# Baseline characteristics among 101,407 people with myocardial infarction over nine-year time period: a population-based study using primary care data

**DOI:** 10.1101/2024.07.17.24310434

**Authors:** Corneliu T Arsene

## Abstract

CardioVascular (CV) Disease (CVD) accounts for 25% of all UK deaths [1-4]. Before developing and applying new or existing CV risk prediction tools [5-22] for prediction of incident CVD events, it is important to have a description of the baseline characteristics of the patients who experienced a myocardial Infarction (MI) event. This paper presents the baseline characteristics of a cohort of patients who experienced a first MI between 1st of January 2006 and 31st of December 2014 in England, UK, age equal or greater than 35 years

## Methods

It is used the Aurum database from the Clinical Practice Research Datalink (CPRD), which includes patients from primary care, linked with practice level Index of Multiple Deprivation (IMD) data to ascertain patients diagnosed for the first time in their lives with MI between 1st of January 2006 and 31st of December 2014 in England, UK, age equal or greater than 35 years. It is reported the baseline characteristics of this population: biological parameters, comorbidities and medications, at the time of the first MI event which took place within the considered time window. It is also reported age, gender, follow-up time (i.e. defined as duration from first MI until censoring defined as the earliest date of end of study 31^st^ of December 2014, date of no survival, transferred out date and last collection date), time to no survival after first MI (i.e. defined as time from first MI until time of no survival taken place before the end of study of 31^st^ of December 2014) and deprivation information.

## Results

For the period of study of 1st of January 2006 to 31st of December 2014, a total of 101,407 patients met the inclusion criteria: 63,939 people were male patients (63%) and 37,468 were female patients (37%), age (mean ± standard deviation) at diagnosis was lower for male patients (66.81 ± 13.19) compared to female patients (74.83 ± 13.01). Within the above period of time of 9 years (i.e. 1st of January 2006 to 31st of December 2014), there were 14,192 who had a first MI and did not survive beyond the date of 31st of December 2014 because of cardiovascular reasons, or 14% of the cohort: from this subgroup (i.e. 14,192) more than 85% were older than 70 years old (i.e. 12,101). The MI follow-up in years (mean ± standard deviation) was 3.24 ± 2.52 years when using the end date of 31st of December 2014. Time to no survival after first MI was a little bit longer for male patients (3.83 ± 3.29) than for female patients (3.39 ± 3.09) when using the end date of 31st of December 2014: however, if the cohort of 101,407 patients would be followed for another 5 years beyond the above end date of 31st of December 2014 (i.e. 31st of December 2019), then the time to no survival after first MI would show a more clear distinction with 6.21 ± 3.87 for male patients and 5.20 ± 3.85 for female patients.

Patients were homogeneously distributed in terms of social deprivation (IMD quintiles): 20% quintile 1-least deprived, 20.4% quintile 2, 19.9% quintile 3, 19.6% quintile 4, 19.8% quintile 5-most deprived.

In terms of Body Mass Index (BMI), 1.5% was underweight, 21% normal weight, 28.5% overweight, 21% obese and 26% had a missing value.

The biological parameters consisted of: Diastolic Blood Pressure (mean ± standard deviation) which was 78.55 ± 11.27 mm Hg for male patients and 78.22 ± 11.69 for female patients, Systolic Blood Pressure was 127.97 ± 16.31 mm Hg for male patients and 131.98 ± 17.84 mm Hg for female patients, Plasma cholesterol was 4.33 ± 1.07 mmol/L for female patients and 3.84 ± 0.99 mmol/L for male patients, Plasma triglyceride was 1.50 ±0.99 mmol/L for male patients and 1.45±0.91 mmol/L for female patients, and High Density Lipoprotein (HDL)-cholesterol was 1.15 ± 0.33 mmol/L for male patients and 1.43 ± 0.38 mmol/L for female patients.

For comorbidities: 10.62% of the cohort had a record of stroke or transient ischemic attack, 6.91% heart failure, 41.33% hypertension, 19.27% chronic kidney disease, 31.87% coronary heart disease, 19.20% diabetes, 20.68% hyperlipidemia, 36.40% family history of CVD, 11.66% cardiovascular procedures, 13.24% chronic pulmonary disease, 9.53% atrial fibrillation, 1.11% heart valve disease, 6.74% peripheral vascular disease, 0.45% pericardial disease, 7.91% hypothyroidism, 17.41% angina, 3.23% rheumatoid arthritis, 0.54% cardiomyopathy. In terms of medications: 48.09% of the cohort were treated with angiotensin-converting enzyme inhibitors (ACEI) and angiotensin-II receptor blockers (ARB), 50.01% lipid-regulating drugs (statins), 15.56% alpha-blockers, 7.11% anticoagulants, 49.11% antiplatelets, 42.62% beta-blockers, 40.11% calcium-channel-blockers, 16.81% diabetes mellitus treatment, 28.88% antianginal, 46.08% diuretics and 71.28% any antihypertensives including centrally-acting (e.g. Methoserpidine), Vasodilators or combined (e.g. beta/thiazide/potassium).

## Conclusions

This paper identified the baseline characteristics (biological, comorbidities, medication and deprivation) of an MI cohort in England, UK between 2006 and 2014 and based on the CPRD Aurum database. From the comorbidities the highest was hypertension, while for the medications was the antihypertensives. It was also noticed that male patients tend to survive a little bit more (3.83 ± 3.29) than female patients (3.39 ± 3.09) after a first MI within the considered time window of 9 years (i.e. 1st of January 2006 to 31st of December 2014), or 6.21 ± 3.87 for male patients and 5.20 ± 3.85 for female patients if considering a longer period of time of 14 years (i.e. 1st of January 2006 to 31st of December 2019). Fig 1 shows also the entire MI cohort (i.e. 101,407 patients) by age groups: 946 patients for lower than 40 years old, 7656 patients for 40 to 49 age interval, 16526 patients for 50 to 59 age interval, 22678 patients for 60 to 69 age interval and 53601 patients equal or above 70 years old.

**Fig. 1.**
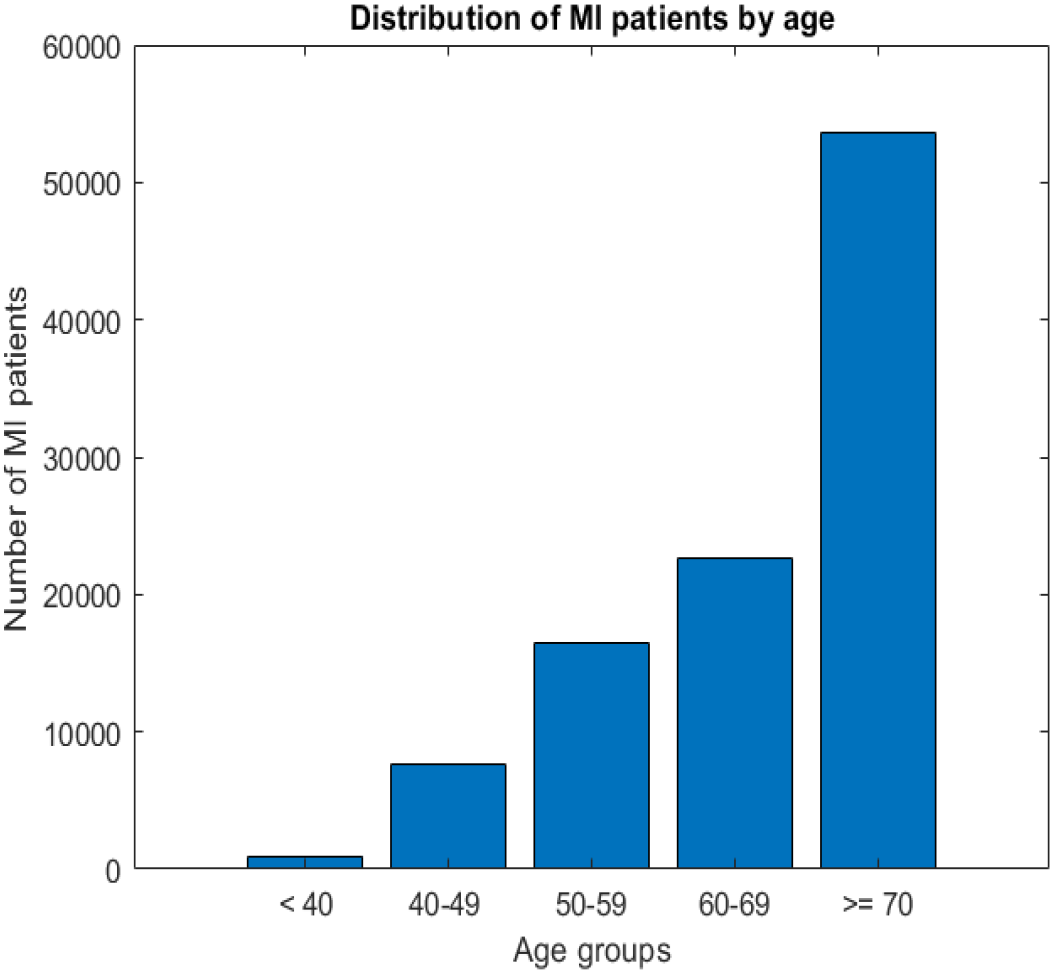
Distribution of MI patients by age groups: 946 patients < 40 age, 7656 patients for 40 to 49 age interval, 16526 patients for 50 to 59, 22678 patients for 60 to 69, 53601 patients for equal or above 70 years old.

## Data Availability

Data is not available.

## Acknowledgment

The author would like to thank to the Heart Research UK – Translation Research Project Grants for supporting this work. The author would like to thank to Dr Salwa Zghebi and Prof Mamas Mamas for the medical and the pharmacological inputs, and also to Prof. Evangelos Kontopantelis and Dr Rosa Parisi for their support.

## Details of ethics approval

This study is based in part on data from the Clinical Practice Research Datalink (CPRD) obtained under licence from the UK Medicines and Healthcare products Regulatory Agency. The data is provided by patients and collected by the NHS as part of their care and support. The interpretation and conclusions contained in this study are those of the author/s alone. Copyright ©[2024], re-used with the permission of The Health & Social Care Information Centre. All rights reserved. The study protocol was approved by the CPRD’s Independent Scientific Advisory Committee (protocol number 20_177). Generic ethical approval for observational research using CPRD with approval from ISAC has been granted by a Health Research Authority (HRA) Research Ethics Committee (East Midlands—Derby, REC reference number 05/MRE04/87).

## References

[1] Gale C., “Acute coronary syndrome in adults: Scope of the problem in the UK”, Br J Cardiol., 24:3–9, 2017.

[2] Wright FL, Townsend N, Greenland M, et al., “Long-term trends in population-based hospitalisation rates for myocardial infarction in England: a national database study of 3.5 million admissions”, 1968–2016. J Epidemiol Community Health, 76(1):45–52, 2022.

[3] Smolina K, Wright FL, Rayner M, et al., “Incidence and 30-day case fatality for acute myocardial infarction in England in 2010: national-linked database study”, The European Journal of Public Health, 22(6):848–53, 2012.

[4] Bhatnagar P, Wickramasinghe K, Wilkins E, et al., “Trends in the epidemiology of cardiovascular, disease in the UK”, Heart, 102(24):1945–52, 2016.

[5] Hippisley-Cox, J., “Derivation and validation of QRISK, a new cardiovascular disease risk score for the United Kingdom: prospective open cohort study”, BMJ, 335:136, 2007.

[6] Hippisley-Cox, J., Coupland, C., Brindle, P., “Development and validation of QRISK3 risk prediction algorithms to estimate future risk of cardiovascular disease: prospective cohort study”, BMJ, 357, 2017.

[7] van Staa, T-P, Gulliford, M., Ng, E.S.-W, Goldacre, B., Smeeth, L., “Prediction of cardiovascular risk using Framingham, ASSIGN and QRISK2: How well do they predict individual rather than population risk?”, PLOS ONE, 2014.

[8] Woodward M, Brindle P, Tunstall-Pedoe H, “SIGN Group on Risk Estimation. Adding social deprivation and family history to cardiovascular risk assessment: the ASSIGN score from the Scottish Heart Health Extended Cohort (SHHEC)”, Heart, 2007, 93:172–176.

[9] Conroy, R.M., Pyorala, K., Fitzgerald, A.E., Sans, S., Menotti, A., De Backer, G., De Bacquer, D., Ducimetiere, P., Jousilahti, P., Keil, U., Njolstad, I., “Estimation of ten-year risk of fatal cardiovascular disease in Europe: the SCORE project”, Eur. Heart J., 2003, 24:987–1003.

[10] Nilsen, A., Hanssen, T.A., Lappegard, K.T., Eggen, A.E., Lochen, M-L., Selmer, R.M., Njolstad, I., Wilsgaard, T., Hopstock, L.A., “Change in cardiovascular risk assessment tool and updated Norwegian guidelines for cardiovascular disease in primary prevention increase the population at risk: the Tromso Study 2015-2016”, vol.8(2), BMJ OpenHeart, 2021.

[11] Zghebi, S.S., Mamas, M.A, Ashcroft, D.M., Rutter, M.K., VanMarwijk, H., Salisbury, C., Mallen, C.D., Chew-Graham, C.A., Qureshi, N., Weng, S.F., Holt, T., Buchan, I., Peek, N., Giles, S., Reeves, D., Kontopantelis, E. “Assessing the severity of cardiovascular disease in 213 088 patients with coronary heart disease: a retrospective cohort study”, Open Heart, 8, 2021.

[12] Smolina, K, Lucy Wright, F., Rayner, M., Goldacre, M., “Long-term survival and recurrence after acute myocardial infarction in England, 2004 to 2010”, vol.5(4), Circulation: Cardiovascular Quality and Outcomes, 2012.

[13] Stromback, U., Vikman, I., Lundblad, D., Lundqvist, R., Engstrom, A., “The second myocardial infarction: higher risk factor burden and earlier second myocardial infarction women compared with men. The Northern Sweden MONICA study”, European Journal of Cardiovascular Nursing, vol.16(5), 418–424, 2017.

[14] Lisboa, PJL, Etchells, T., Jarman, I, Arsene, CTC, Aung, M.S.H. Eleuteri, A. Taktak, A.F.G. Ambrogi, F. Boracchi, P. Biganzoli, E., “Partial Logistic Artificial Neural Network for Competing Risks Regularized With Automatic Relevance Determination”, IEEE Transactions on Neural Networks, Vol 20, Issue 9, 1403–1416, 2009.

[15] Arsene, C.T.C., & Lisboa, P.J.C., “Artificial neural networks used in the survival analysis of breast cancer patients: a node negative study”, in Perspectives in Outcome Prediction in Cancer. Amsterdam: Elsevier Science Publ, Editors: A.F.G. Taktak and A.C. Fisher; ISBN, 2007.

[16] Arsene, C.T.C., P. Lisboa, E. Biganzoli, “Model Selection with PLANN-CR-ARD”, J. Cabestany, I. Rojas, and G. Joya (Eds.): IWANN 2011, Part II, Lecture Notes in Computer Science (LNCS) 6692, pp. 210–219, 2011, ©Springer-Verlag Berlin Heidelberg, 2011, 2011.

[17] Arsene, C.T.C., P.J. Lisboa, “Bayesian Neural Network with and without compensation mechanism for competing risks”, Proceedings of the International Joint Conference on Neural Networks (IJCNN), Brisbane, Australia, 2012.

[18] Arsene, C.T.C., P.J. Lisboa, “PLANN-CR-ARD model predictions and Non-parametric estimates with Confidence Intervals”, Proceedings of the International Joint Conference on Neural Networks (IJCNN), San Jose, California, 2011.

[19] Arsene, C.T.C., “Partial Logistic Artificial Neural Network with Automatic Relevance Determination and Markov Chain More Carlo methods applied in medical survival studies”, IEEE Computational Intelligence in Bioinformatics and Computational Biology (CIBCB), Chiang Mai, Thailand, 2016.

[20] Arsene, C.T.C., Lisboa, P.J.G., Borrachi, P., Biganzoli, E. and Aung, M.S.H., “Bayesian Neural Networks for Competing Risks with Covariates”, The Third International Conference in Advances in Medical, Signal and Information Processing, MEDSIP 2006, July 2006, 2006.

[21] Arsene, C.T.C., Lisboa, P.J., “A predictive and prognostic medical study for Primary Billiary Cirrhosis by using a Bayesian Neural Network”, E-Health and Bioengineering conference, EHB 2011, 2011.

[22] Lisboa, PJG, Wong, H., Harris, P, Swindell, R., “A Bayesian neural network approach for modeling censored data with an application to prognosis after surgery for breast cancer”, Artificial Intelligence in Medicine, vol.28(1), 1–25, 2003.

